# Mosaic loss of chromosome Y and coronary atherosclerosis in men: insights into sex differences in cardiovascular risk from the SCAPIS study

**DOI:** 10.1101/2025.07.10.25331326

**Authors:** Josefin Bjurling, Andrei Malinovschi, Tomas Jernberg, Carl Johan Östgren, Gunnar Engström, Anders Blomberg, Anders Gummesson, Laxmipriya Ramesh, Ammar Zaghlool, Tove Fall, Marcel den Hoed, Jonatan Halvardson, Lars A. Forsberg

## Abstract

**Background:** Atherosclerosis predominantly affects men and is the underlying cause of coronary artery disease (CAD) and myocardial infarction (MI). Recent studies show that mosaic loss of chromosome Y (LOY) in blood is associated with all major causes of death, including CAD and MI. However, the relation between LOY and subclinical atherosclerosis remains unclear. This study aims to investigate associations between the male-specific LOY and atherosclerosis.

**Methods:** To examine associations between hematopoietic LOY and atherosclerosis prevalence, we analyzed imaging summary information and genotyping data from 30,000 participants (∼50% men) in the Swedish CArdioPulmonary bioImage Study (SCAPIS). The level of LOY in whole blood DNA samples from 12,390 male participants was estimated from SNP-array data using established bioinformatics methods. Thereafter, men were divided into three groups based on the level of Y loss in blood: no detectable LOY, low LOY (£10%), and substantial LOY (>10%). Low and substantial chromosome Y loss was modelled in multivariable adjusted logistic regression analyses to assess associations with male atherosclerosis, measured by coronary computed tomography angiography (CCTA) and carotid ultrasound.

**Results:** We report that substantial LOY in blood leukocytes is associated with the prevalence of coronary atherosclerosis (OR=1.48, 95%CI=1.07-2.06, p=0.02) as well as the occurrence of substantial coronary stenosis (OR=1.56, 95% CI=1.05-2.25, p=0.02). We further show that men with substantial LOY exhibit a higher number of involved segments (OR=1.2, 95%CI=1.004-1.439, p=0.047) and an increased accumulation of calcified plaques (OR=1.56, 95%CI=1.08-2.37, p=0.02). In this dataset, the occurrence of carotid plaques was not associated with LOY after multivariable adjustment (OR=1.17, 95%CI=0.86-1.62, p=0.33).

**Conclusions:** We report that LOY in circulating blood leukocytes is associated with coronary atherosclerosis in men. These results contribute to understanding the male predominance in atherosclerosis and related cardiovascular outcomes. We hypothesize that LOY-driven atherosclerosis represents a mechanistic link in the previously described associations between LOY in blood leukocytes and major cardiovascular events in men.

## Introduction

Atherosclerosis and its clinical manifestations, including coronary artery disease (CAD) and myocardial infarction (MI), are the leading causes of morbidity and mortality globally^1,2^. The prevalence of atherosclerosis is higher among men, who also have a disease onset on average 12 years earlier than women^3–5^. Environmental and cardiovascular risk factors have been proposed to explain the diverging prevalence in men and women^4^. However, recent studies observed only slightly attenuated risk estimates when correcting for these variables^3,4^. Since the cause of the sex discrepancy remains unknown, we examined whether the male-specific condition, mosaic loss of chromosome Y (LOY), could contribute to the higher burden of atherosclerosis in men.

Hematological LOY is a known risk factor for cardiovascular disease (CVD) in men^6–11^, including atherosclerotic diseases like CAD^12^ and MI^12^. LOY was also associated with functional outcomes after ischemic stroke^13^. Although a slight increase in atheroma size has been noted in men with LOY in a male atherosclerosis cohort^6^, the connection between LOY and atherosclerosis prevalence has not yet been assessed. A higher degree of LOY is observed in older men, current smokers, and/or those who have an underlying genetic predisposition for genomic instability^14–16^. A study conducted on the UK Biobank cohort estimated 20% of male participants to be affected by hematological LOY^16^. However, data from single-cell studies suggest that all men carry cells without the Y chromosome^16–18^. Besides CVD, other diseases such as Alzheimer’s disease^19–21^, lung diseases like idiopathic pulmonary fibrosis^22,23^, cancers^24–28^, and infections^22,28–30^ have also been associated with hematological LOY.

The connection between LOY in blood and disease risk in other organs has started to emerge from studies demonstrating an altered epigenetic and transcriptional landscape in infiltrating immune cells with LOY, resulting in impaired cellular function^17,18,31–34^. Moreover, LOY in blood has been shown to cause fibrosis of the mouse heart, lung, and other organs, through a TGFb1-driven mechanism^35^. This profibrotic disease mechanism was recently validated in humans as LOY in tissue-infiltrating leukocytes was shown to have enhanced TGFb signaling in men with pulmonary fibrosis^23^. In line with this, cardiac disease outcomes in men with LOY are characterized by a fibrotic phenotype^9,12,36^.

In this study, we examine the sex difference of atherosclerosis and whether hematological LOY is associated with the occurrence and burden of coronary atherosclerosis and/or carotid plaques using extensive imaging data from participants in the Swedish CArdioPulmonary bioImage Study (SCAPIS).

## Methods

### SCAPIS study participants

A detailed description of the prospective SCAPIS cohort and the cardiovascular examinations performed on SCAPIS participants has previously been provided by Bergström et al.^5,37^ Briefly, 30,154 individuals aged 50-64 years (50% men) from six different sites in Sweden were enrolled between 2013 and 2018. Individuals who withdrew their participation or did not provide consent were excluded from the present study, resulting in a dataset of 30,026 participants. At the initial visit, core examinations were performed on all participants. This included self-completed questionnaires, imaging and functional measures of the cardiovascular and pulmonary systems, collection of biological samples for biochemistry analyses, and array-based whole-genome genotyping. Coronary arteries were examined using Coronary Computed Tomography Angiography (CCTA), and results were reported by dividing the arteries into 18 anatomical segments according to guidelines from the Society of Cardiovascular Computed Tomography^38^. A dual-source scanner equipped with a Stellar Detector (Somatom Definition Flash, Siemens Medical Solutions) was utilized to perform the CCTA on all participants without contraindications to contrast media. Examination of the carotid arteries was performed using high-resolution ultrasound. SCAPIS has been approved as a multicentre study by the ethics committee of Umeå University (Dnr 2010-228-31M) and complies with the Declaration of Helsinki. The present study was approved by the Swedish Ethical Review Authority (Dnr 2023-00991-01). All participants provided written informed consent to participate; of these, 29,433 participants gave consent for genetic analyses.

### Estimation of LOY in blood leukocytes

DNA from whole blood was genotyped using a customized version of the Illumina GSA-MDv3 genotyping array^39^. After preprocessing, DNA-array data from 29,356 SCAPIS participants in 10 batches remained for analyses. GenomeStudio (v2.0) was used to re-cluster only the male participants in each batch and to generate log R ratios (LRR) and B allele frequencies (BAF) for the 5,026 probes on the Y chromosome. Thereafter, LOY was estimated using two different approaches for comparison. First, the MoChA package was used to call LOY based on measurements from the pseudoautosomal (PAR) 1 region. Briefly, VCF files containing LRR and BAF values were constructed for probes on chromosomes Y (including PAR1), X, and 10. MoChA was then run using the procedure outlined in https://github.com/freeseek/mocha. LOY values were called using the code outlined under “*Generate list of samples with mosaic loss of chromosome Y (mLOY or LOY*)” ^40,41^. Secondly, LOY was called by calculating the median of the log R ratios for 4,148 probes in the male-specific region of chromosome Y (mLRRY) in R (v 4.3.1), as described before ^24^. The LOY estimates from MoChA were used for all statistical tests.

### Inclusion criterion

Two men with differing LOY estimates from two DNA array runs were removed from the final dataset. Next, the mLRRY and MoChA LOY estimates were validated in a pairwise fashion and seven participants with deviating measures from the two methods were excluded (Supplement Figure 1). BAF values for PAR1 probes were examined for the excluded samples, which revealed that the deviation of LOY measures was due to the occurrence of blood cells with 47,XXY or 47,XYY karyotype (Supplement Figure 2). Individuals with successful CCTA measures in four proximal coronary segments and no coronary intervention or no previous MI according to CCTA measures, self-reported questionnaires, and medical records were used for further analyses (n =25,079)^3,5^.

### Data curation

All data processing was carried out using R v 4.3.1. Summary variables from the CCTA analysis were extracted and used as outcomes. The primary outcome was atherosclerosis measured in any coronary segment. The severity of atherosclerosis was further explored using variables for any stenosis >50% in any coronary segment, segment involvement score (SIS), and the summed coronary artery calcium (CAC) score according to Agatston^42^ from the three coronary arteries. Additionally, carotid atherosclerosis was investigated, setting the variable as no plaque or occurrence of plaque (either one or two vessels). Variables for data processing and statistical analyses were age in years, smoking (non-smoker, previous, and current), total cholesterol (mmol/L), body mass index (BMI), diastolic blood pressure (DBP), systolic blood pressure (SBP), and diabetes. The data extraction variables are provided in Supplement Table 1. The BMI variable was divided into four levels: normal (18.5-24.9), underweight (<18.5), overweight (b25), and obese (b30), with normal set as the reference. Variables for diastolic and systolic blood pressure were changed into factor levels according to the hypertension stages from the American Heart Foundation^43^. Participants glycemic status was classified as normoglycemic, presenting with pre-diabetes (HbA1c >42 mmol/mol and < 48 mmol/mol or impaired fasting glucose) or being diagnosed with diabetes mellitus (previously known diabetes mellitus or new diabetes mellitus diagnosis). Baseline characteristics are presented in Table 1 for the SCAPIS participants included in the present study.

**Table 1.**
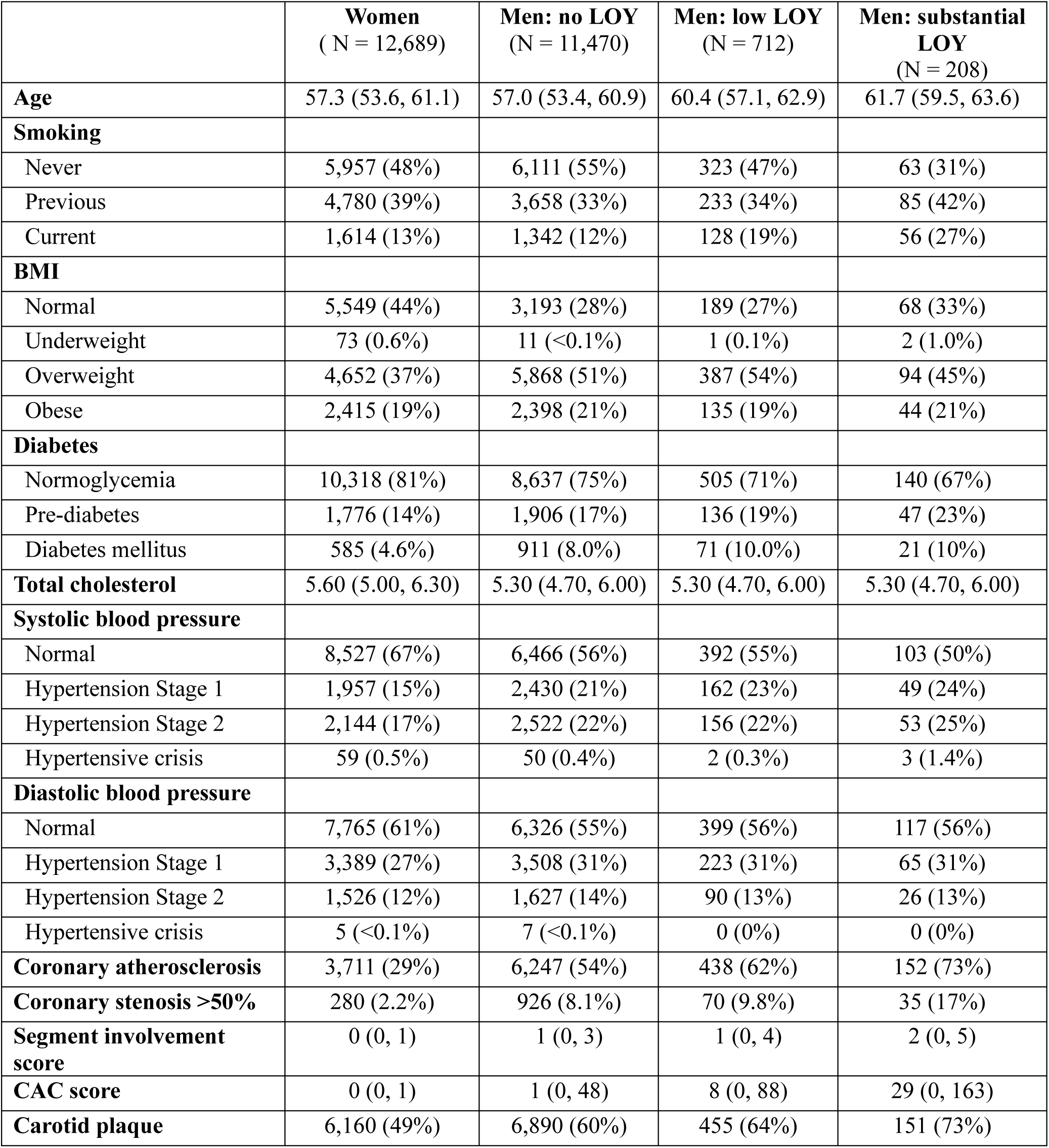
Baseline characteristics for SCAPIS participants. Comparison of the baseline parameters presented for women, men with no LOY, men with low LOY (£10%), and men with substantial LOY (>10%) separately. LOY = mosaic loss of chromosome Y, sub. = substantial, BMI = body mass index, CAC = coronary artery calcium, IQR = interquartile range. Variables are presented as median (IQR) or number (%).

### Statistical analyses

A categorical variable for LOY was created by dividing men into three groups based on clonal fractions of Y loss in blood (no detectable LOY (no LOY, n = 11,470), low LOY (<10%, n = 712), and substantial LOY (>10%, n = 208) (Supplement Figure 3). Analyses exploring sex differences in coronary atherosclerosis and carotid plaques were conducted with data from 12,390 men and 12,689 women. Analyses assessing associations between LOY and atherosclerosis and related traits were performed using data from the male study population. The statistical analyses were conducted using R (v 4.3.1.). To assess the relationship between LOY and the number of segments involved or CAC score, logistic regression analyses were performed with the glm.nb function from the MASS package (v 7.3.60). The analyses for the remaining outcomes, performed to explore the sex difference as well as differences between men with and without LOY, were conducted with the glm function from the stats package (v 4.3.1) using the binomial family argument. In all statistical analyses, the atherosclerosis variables were set as the dependent variable, and all tests were adjusted for age, smoking, BMI, diabetes, total cholesterol, DBP, and SBP (Table 1).

## Results

### A profound sex difference in atherosclerosis

Atherosclerosis prevalence was first explored between 12,390 men and 12,689 women with a successful CCTA and no prior intervention or MI (Supplement Figure 4A) using multivariable logistic regression analyses. Consistent with previous studies, these analyses demonstrated a male predominance, and that men overall had a three-fold increased prevalence of coronary atherosclerosis (adjusted odds ratio (OR) = 3.22, 95% confidence interval (CI) = 3.04-3.41, p <0.0001) (Figure 1A, Supplement Table 2) and a 1.7-fold increased prevalence of carotid plaques (OR=1.7, CI=1.61-1.79, p <0.0001) (Figure 1A, Supplement Table 3) compared with women.

**Figure 1.**
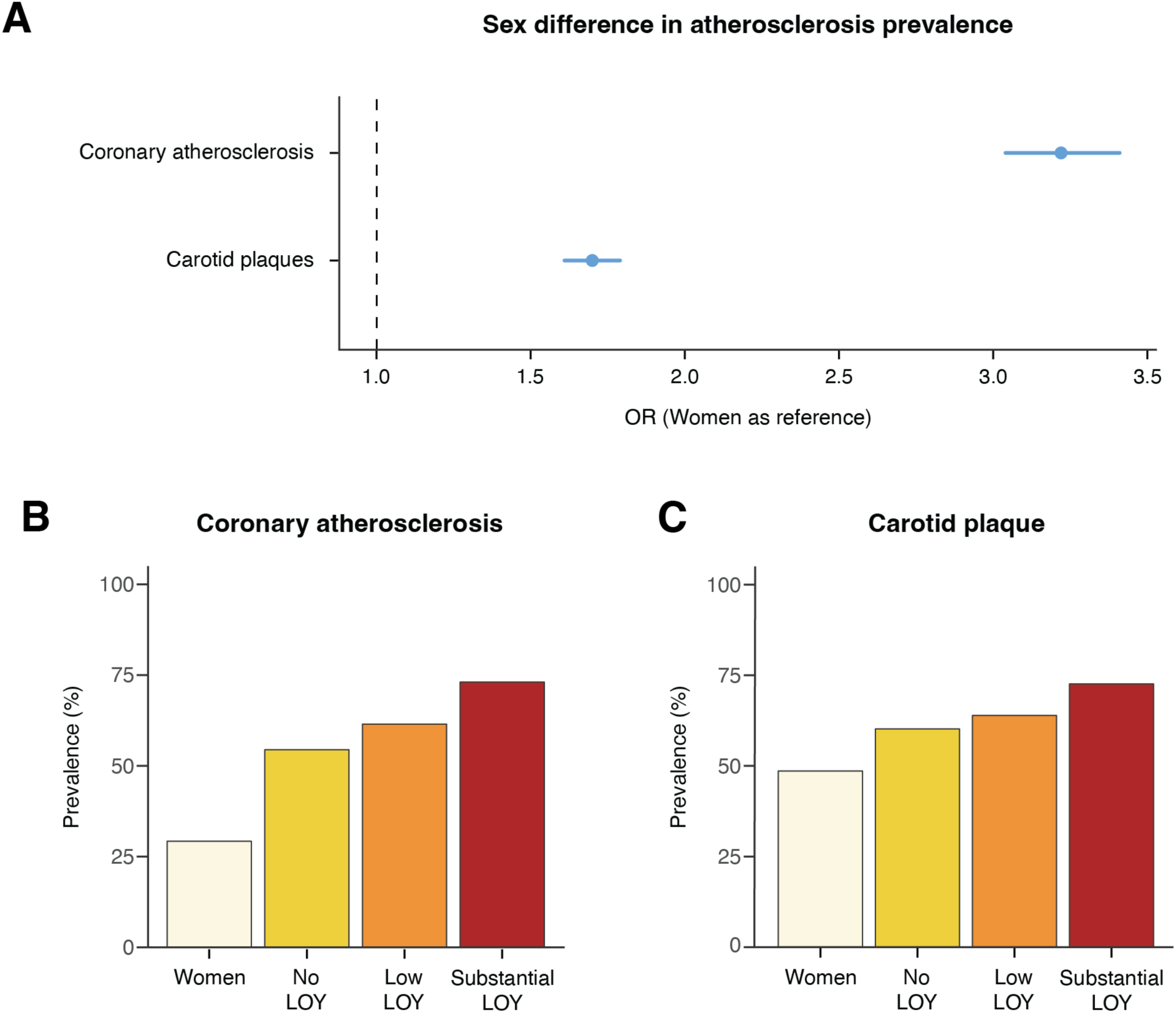
Sex difference in prevalence of atherosclerosis. **A)** Forest plot with the results from the analyses investigating the prevalence of coronary and carotid atherosclerosis in men and women. The analyses were conducted on all SCAPIS participants with valid CCTA readouts. Adjusted odds ratios (OR) and 95% confidence intervals are marked as dots and lines, respectively. The dashed line at OR = 1 indicates no difference between men and women. **B**) Comparison of the observed prevalence of coronary atherosclerosis between women (n = 12,689) and men stratified by their level of LOY in blood: no detectable LOY (no LOY, n = 11,470), £10% LOY (low LOY, n = 712), and >10% LOY (substantial LOY, n = 208). **C**) Comparison of the observed carotid plaque prevalence between the same groups as stated in B.

### LOY in blood associated with coronary atherosclerosis in men

Atherosclerosis in relation to hematological LOY was examined among 12,390 men who remained in the final dataset after data processing (Supplement Figure 4B). The prevalence of coronary atherosclerosis and carotid plaques was observed to be higher in men with LOY in blood compared to other men in unadjusted comparisons (Figure 1B and 1C). We performed multivariable adjusted logistic regression analyses to examine if the association remained after adjusting for relevant confounders. From these analyses, we found that men with substantial LOY display more atherosclerosis in coronary arteries compared with men without LOY (OR = 1.48, 95% CI = 1.08-2.07, p=0.02) (Figure 2, Table 2). In contrast, LOY in blood was not significantly associated with the presence of carotid plaques after multivariable adjustment (Figure 2, Supplement Table 7). There was no statistical difference in the prevalence of either outcome between men with low LOY compared with men with no detectable LOY after multivariable adjustment (Figure 2, Table 2, Supplement Table 7). Following this, the severity of male coronary atherosclerosis in relation to LOY was examined. First, we show that lumen narrowing, as measured by exhibiting any coronary stenosis >50%, was associated with substantial LOY (OR=1.56, CI=1.05-2.26, p=0.02) (Figure 2, Supplement Table 4). Second, the extent of coronary atherosclerosis, estimated by the number of involved segments by the SIS variable, was greater in men with substantial LOY (OR=1.2, CI=1.0035-1.44, p=0.047) (Figure 2, Supplement Figure 5, Supplement Table 5). And third, men with LOY in >10% of peripheral blood cells had a higher summed CAC score, estimating the amount of calcium in three major coronary arteries, compared to men without detectable LOY (OR=1.56, CI=1.08-2.37, p=0.02) (Figure 2, Supplement Table 6). In contrast to men with substantial LOY, men with detectable, but lower levels of LOY mosaicism, did not display enhanced coronary atherosclerosis by any of these measures (Figure 2, Supplement Tables 4-6). Consistent with previous reports, the prevalence of LOY increased with advancing age in male SCAPIS participants (Supplement Figure 6). No significant associations were observed for any atherosclerotic outcomes in sensitivity analyses including only non-smoking male participants (n=6497) (Supplement Figure 4B, Supplement Table 8). Overall, our results indicate that a substantial clonal fraction of blood cells without the Y chromosome is associated with increased prevalence and severity of coronary atherosclerosis in men.

**Figure 2.**
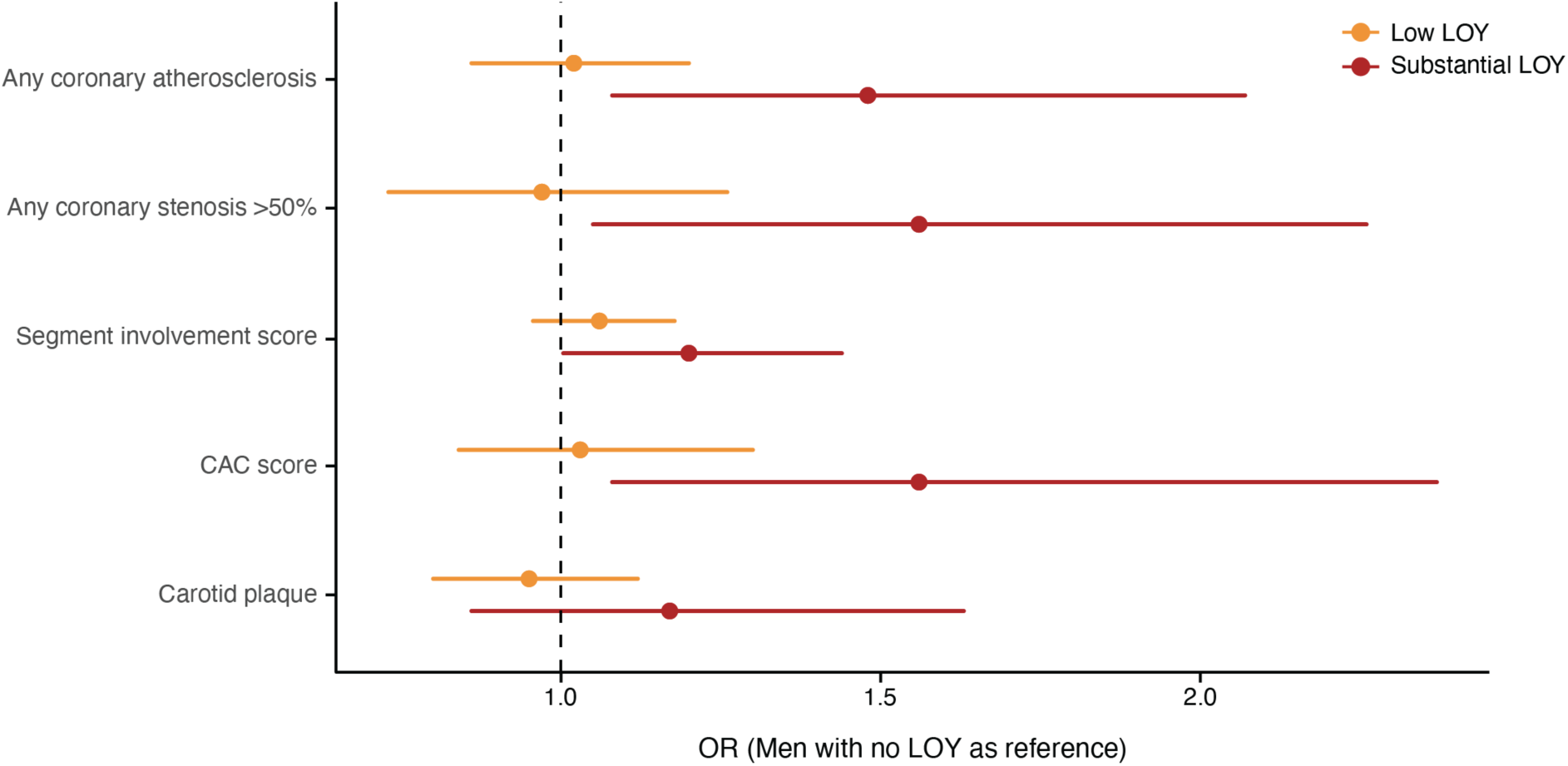
Mosaic loss of chromosome Y in circulating blood leukocytes is associated with coronary atherosclerosis in men. Results from multivariable logistic regression analyses are displayed using a Forest plot showing the difference in atherosclerosis outcomes in men with LOY in low and substantial clonal fractions. The comparisons between men with no detectable LOY (n = 11,470) and men with low LOY (n = 712) are presented in orange, while the results comparing men with no detectable LOY and men with substantial LOY (n = 208) are presented in red. Adjusted odds ratios (OR) are displayed as dots and 95% confidence intervals as lines in the plot. The dashed line indicates no difference between the investigated groups.

**Table 2.**
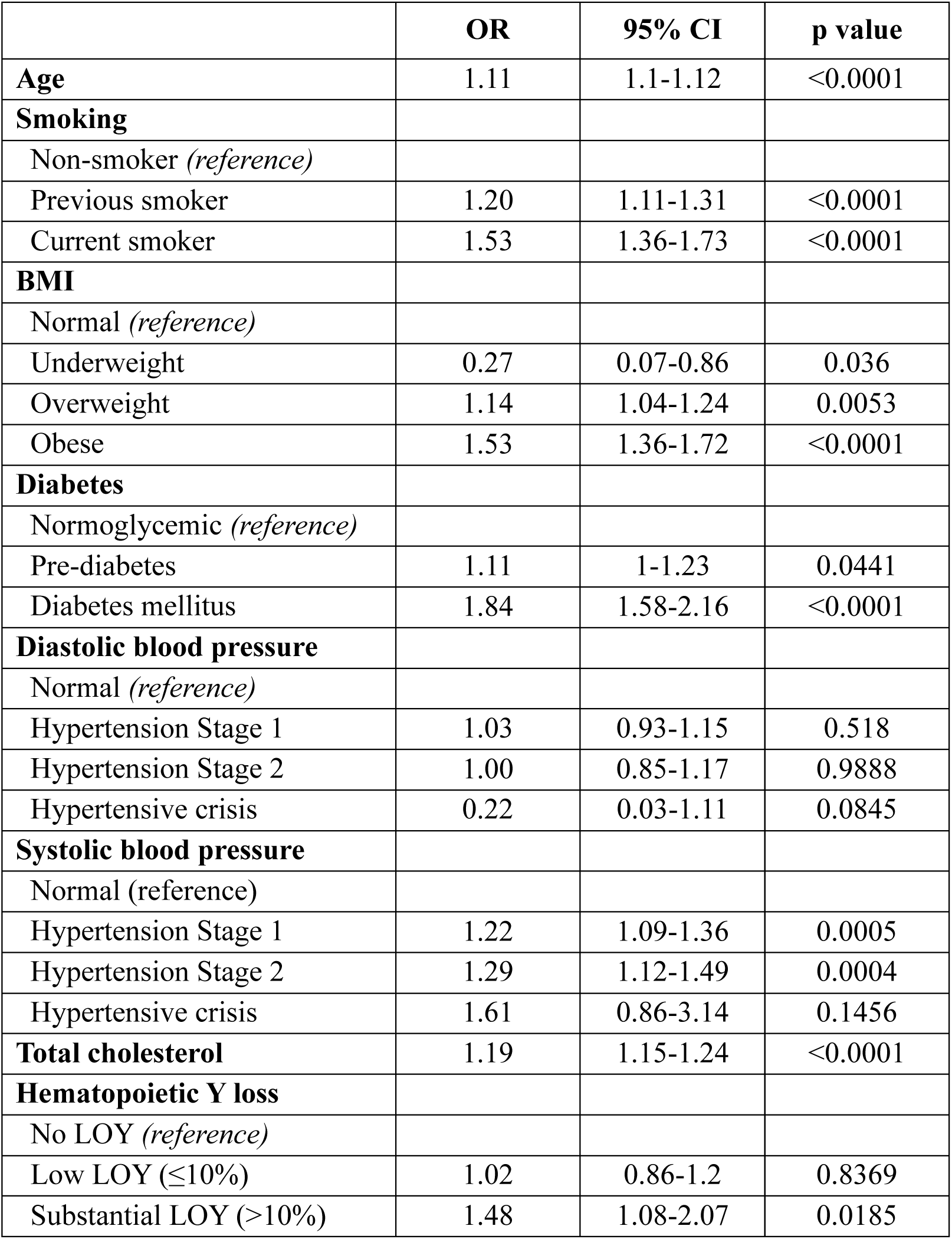
Multivariable logistic regression analysis of coronary atherosclerosis in relation to hematological LOY in men. Multivariable adjusted logistic regression analysis testing differences in prevalence of any coronary atherosclerosis between men with LOY and men with no detectable LOY. The model is adjusted for age, smoking, BMI, diabetes, diastolic and systolic blood pressure and total cholesterol. OR = odds ratio, CI = confidence interval, BMI = body mass index, LOY = mosaic loss of chromosome Y.

## Discussion

Atherosclerosis exhibits a well-established male predominance and constitutes a primary underlying cause of MI and nearly 75% of CVD deaths^1^. Previous studies of the SCAPIS cohort describe a high and male-prevalent subclinical atherosclerosis^3,5^. Here we present results suggesting that Y chromosome loss in blood contributes to the atherosclerotic burden of men. Previous studies of clinical and epidemiological cohorts showed associations with LOY in circulating leukocytes for major cardiovascular events in men^6,8–11,35^, including MI mortality in men with CAD^12^. In the latter study, Weyrich *et al.* studied the association between LOY in peripheral blood and cardiovascular mortality among 1,698 male patients undergoing coronary angiography. It should be noted that the methods applied in that study did not directly quantify the extent or characteristics of atherosclerosis within the coronary arteries. Rather, the study relied on angiographic identification of CAD and subsequent clinical outcomes, without direct imaging or quantification of atherosclerotic plaque burden or composition. Thus, while their findings robustly link and replicate associations between LOY in blood with increased cardiovascular and all-cause mortality^12^, the study leaves unresolved whether LOY is associated with the presence, burden, or phenotype of atherosclerotic plaques per se.

To address this gap, we examined CCTA data collected within the SCAPIS study to directly assess atherosclerosis in the coronary vasculature. Compared with conventional coronary angiography, CCTA offers high sensitivity and specificity for detecting coronary atherosclerosis, enabling direct measurements of the presence and extent of coronary atherosclerosis, as well as coronary calcification. Utilizing these direct measures of atherosclerosis from >12,000 men, 50-64 years of age, from the general Swedish population, our results corroborate and extend previous findings by showing a greater prevalence of coronary atherosclerosis among men with LOY in blood leukocytes. In contrast to the clear association between coronary atherosclerosis and LOY, we found no association with atherogenic traits of the carotid arteries. This discrepancy is in line with the male predominance being more evident in coronary compared with carotid atherosclerosis^3^ and provides a hint towards the molecular mechanisms that may play a role. Coronary atherosclerosis, on the one hand, is characterised by the accumulation of lipids, inflammatory cells – especially macrophages – and fibrous tissues within the intimal layer of the coronary arteries. It is fuelled by hyperlipidaemia, endothelial dysfunction, smooth muscle cell migration and proliferation, and matrix remodelling^44^. Hypertrophy of carotid intima-media, on the other hand, is a subclinical marker that may reflect an adaptive response to hypertension and hemodynamic stress^45^. Thus, carotid plaque formation may be driven by hypertension rather than lipid accumulation^45^. Alternatively, the observed differences may be attributable to the varying capacity of the imaging modalities employed; CCTA provides superior accuracy in detecting coronary atherosclerosis compared to the high-resolution ultrasound used for carotid assessments. Future studies might clarify whether these methodological differences are masking a connection between LOY and the occurrence of carotid plaques, which could not be detected in this dataset.

We have previously reported that men in the UK Biobank with LOY in blood display an overall increased risk of death from diseases of the cardiovascular system^35^. In that study, we have also demonstrated that hematopoietic LOY in mice leads to cardiac fibrosis and heart failure mortality, mediated via upregulation of TGFβ-signaling and activation of profibrotic gene programs in macrophages. This mechanistic link was further supported by the observation that pharmacological inhibition of TGFβ could reverse the fibrotic phenotype and restore cardiac function in LOY-mice. Importantly, the profibrotic effects of LOY in blood are not limited to mouse models, as recent studies demonstrate a profibrotic gene signature sensitizing human circulating LOY-monocytes for the TGFβ-signaling pathways^9^ and that LOY is associated with increased risk of being diagnosed with and dying from idiopathic pulmonary fibrosis^22,23^. Briefly, we recently replicated in humans the profibrotic disease mechanism first discovered in mouse models^23^. These findings suggest that LOY in blood may contribute to multi-organ fibrosis and related complications. In the context of atherosclerosis, it is plausible that such LOY-driven fibrogenic pathways alongside inflammation could play a significant role in plaque development, stability, and the progression of vascular disease. Luminal narrowing of the coronary arteries is a key determinant of ischemic heart disease risk, as it impairs myocardial perfusion^46^. Haitjema et al. found a connection between LOY and plaque size occluding >10% of the luminal diameter in a cohort of men with femoral or carotid atherosclerosis^6^. While information required to examine such associations in carotid plaque is not available to us, we observed an association between >50% coronary stenosis and LOY in blood, indicating clinically relevant narrowing of the coronary arteries. Additionally, we found that men with LOY exhibited a higher prevalence of calcified plaques and elevated CAC scores, both of which are established predictors of future cardiovascular events. Taken together, the available data suggest that LOY may contribute to the formation of stable plaques, characterized by substantial calcification and a fibrous cap^47^, which in turn are associated with atherosclerotic diseases such as CAD and MI^12^.

A notable strength of the present study is the use of detailed CCTA imaging, which allows for comprehensive characterization of coronary atherosclerosis across all major vessels. The availability of both CCTA and carotid ultrasound data permitted exploration of LOY associations at different vascular sites. However, limitations such as different methods used for coronary and carotid measurements should be acknowledged. Furthermore, a low number of non-smoking men with substantial LOY in the current dataset prevents robust sensitivity analyses. Additionally, the data currently available in SCAPIS limits our ability to assess the relationship between LOY, atherosclerosis progression, and incident cardiovascular events.

In conclusion, this study is the first to report a direct association between coronary atherosclerosis and mosaic loss of chromosome Y in blood. These findings, together with previous evidence^9,12,35^, support the hypothesis that LOY contributes to the sex difference in adverse cardiovascular outcomes by promoting profibrotic and atherosclerotic processes, providing insight into the higher CVD prevalence among men in the general population.

## Data Availability

The data that support the findings of this study are available from the SCAPIS (Swedish CArdioPulmonary bioImage Study) cohort. Due to legal and ethical restrictions related to participant privacy, SCAPIS data are not publicly available. However, qualified researchers affiliated with a Swedish research institution, or international researchers collaborating with a Swedish institution, may apply for access to the data.

## Acknowledgments

This research has been conducted using the The Swedish CArdioPulmonary bioImage Study (SCAPIS) Resource, under Petition Number 670. The main funding body of SCAPIS is the Swedish Heart and Lung Foundation. The study is also funded by the Knut and Alice Wallenberg Foundation, the Swedish Research Council, VINNOVA (Sweden’s Innovation agency), the University of Gothenburg and Sahlgrenska University Hospital, Karolinska Institutet and Region Stockholm, Linköping University and University Hospital, Lund University and Skåne University Hospital, Umeå University and University Hospital, Uppsala University and University Hospital. We would like to acknowledge the help of Biobank Sweden and the local biobank facilities for their services in handling of biological samples and biobanking. Genotyping was performed by the SNP&SEQ Technology Platform in Uppsala. The facility is part of NGI Sweden and Science for Life Laboratory. The SNP&SEQ Platform is also supported by the Swedish Research Council and the Knut and Alice Wallenberg Foundation. The computations and data handling were made possible by resources from project sens2019512 provided by the Swedish National Infrastructure for Computing (SNIC) at Uppsala Multidisciplinary Center for Advanced Computational Science (UPPMAX), partially funded by the Swedish Research Council through grant agreement no. 2018-05973. We would like to acknowledge the Karolinska Institute Biobank for their services regarding DNA extraction. The storage and analysis of SCAPIS data were enabled by resources in project sens2017-134 provided by the National Academic Infrastructure for Supercomputing in Sweden (NAISS) at UPPMAX, which was funded by the Swedish Research Council (2022-06725).

## Supplemental Materials

Supplemental Tables 1-8

Supplemental Figures 1-6

